# A context-based chatbot surpasses trained radiologists and generic ChatGPT in following the ACR appropriateness guidelines

**DOI:** 10.1101/2023.04.10.23288354

**Authors:** A Rau, S Rau, A Fink, H Tran, C Wilpert, J Nattenmueller, J Neubauer, F Bamberg, M Reisert, MF Russe

**Affiliations:** Department of Diagnostic and Interventional Radiology, Medical Center – University of Freiburg, Faculty of Medicine, University of Freiburg, 79106 Freiburg, Germany; Department of Neuroradiology, Medical Center – University of Freiburg, Faculty of Medicine, University of Freiburg, 79106 Freiburg, Germany; Medical Physics, Department of Diagnostic and Interventional Radiology, Medical Center – University of Freiburg, Faculty of Medicine, University of Freiburg, 79106 Freiburg, Germany; Department of Stereotactic and Functional Neurosurgery, Medical Center - University of Freiburg, Faculty of Medicine, University of Freiburg, 79106 Freiburg, Germany

## Abstract

**Background:** Radiological imaging guidelines are crucial for accurate diagnosis and optimal patient care as they result in standardized procedures and thus reduce inappropriate imaging studies. In the present study, we investigated the potential to support clinical decision-making using an interactive chatbot designed to provide personalized imaging recommendations based on indexed and vectorized American College of Radiology (ACR) appropriateness criteria documents.

**Methods:** We utilized 209 ACR appropriateness criteria documents as specialized knowledge base and employed LlamaIndex and the ChatGPT 3.5-Turbo to create an appropriateness criteria contexted chatbot (accGPT). Fifty clinical case files were used to compare the accGPT’s performance against radiologists at varying experience levels and to generic ChatGPT 3.5 and 4.0.

**Results:** All chatbots reached at least human performance level. For the 50 case files, the accGPT provided a median of 83% (95% CI 82-84) ‘usually appropriate’ recommendations, while radiologists provided a median of 66% (95% CI 62-70). GPT 3.5-Turbo 70% (95% CI 67-73) and GPT 4 79% (95% CI 76-81) correct answers. Consistency was highest for the accGPT with almost perfect Fleiss’ Kappa of 0.82. Further, the chatbots provided substantial time and cost savings, with an average decision time of 5 minutes and a cost of 0.19 Euro for all cases, compared to 50 minutes and 29.99 Euro for radiologists (both p < 0.01).

**Conclusion:** ChatGPT-based algorithms have the potential to substantially improve the decision-making for clinical imaging studies in accordance with ACR guidelines. Specifically, a context-based algorithm performed superior to its generic counterpart, demonstrating the value of tailoring AI solutions to specific healthcare applications.

## Introduction

The increasing demand for accurate and efficient diagnostic imaging in modern healthcare requires standardized criteria for decision-making. The American College of Radiology (ACR) provides such recommendations since 1994 that streamline decision-making processes for referring clinicians ^1^. This allows for optimized patient care, improved diagnostic accuracy and lower radiation exposure, and reduces healthcare costs (https://www.acr.org/Clinical-Resources/ACR-Appropriateness-Criteria). However, variability in clinical routine still persists regarding requirement of imaging itself, modality and need for contrast agent leading to a substantial amount of inappropriate imaging procedures ^2–7^.

Adherence to appropriateness guidelines is highly dependent on clinicians‘ and radiologists’ experience and hampered by a lack of awareness ^4,8–12^. Additionally, the rapid evolution of imaging technology and the progress of clinical evidence necessitate continuous updates to the recommendations, further complicating their usage.

Several clinical-decision support tools were introduced to improve usage of published guidelines (e.g. ESR iGUIDE (https://www.myesr.org/esriguide) or CareSelect® Imaging; (https://www.changehealthcare.com/clinical-decision-support/careselect/imaging)) ^10,13^. These tools have proven valuable in improving the diagnostic management of patients with various clinical conditions, resulting in lower rates of inappropriate examinations ^3,14,15^. However, they require substantial human interaction while potentially losing relevant clinical information as a free-text input is not feasible ^3^.

Artificial intelligence (AI)-based algorithms employing large language models (LLM) can address these limitations by comprehending and interpreting human language. OpenAI introduced ChatGPT to a wide audience in November 2022. This chatbot, specifically trained for conversation, is based on a generative pre-trained transformer and the latest iteration GPT 3.5-Turbo. ChatGPT and especially using GPT 4 (released in March 2023, with at the moment still limited access) were shown to provide substantial medical knowledge being able to pass the USMLE ^16,17^.

ChatGPT enables rapid processing of complex information and its potential applications in clinical radiology routines have been extensively explored and published in preprints. These comprise the preparation of radiological reports ^18^, transferring radiological reports into plain language by simplifying them ^19,20^, or providing clinical decision support on differential diagnoses, diagnostic procedures, final diagnosis, and treatment ^21^.

The community already discusses the potential of ChatGPT to provide recommendations regarding imaging as clinical decision support ^22^ and initial data on breast cancer screening and breast pain are promising ^23^. However, no clinical files but only the respective description of the clinical conditions in the ACR guidelines were presented in the latter study and the comparison to human performance is lacking. Furthermore, ChatGPT is limited to its training data (GPT 3.5-Turbo and GPT 4 trained on data up to September 2021) and thus may not have access to the latest and most specialized knowledge, or might be biased due to the large number of different sources of training data. Therefore, ChatGPT could provide incorrect or incomplete information.

Incorporating specialized knowledge to create an appropriateness criteria contexted GPT (accGPT) should provide more accurate and relevant responses to user queries. To explore this approach, we benchmarked radiologists of varying experience levels and publicly available generic chatbots GPT 3.5-Turbo and GPT 4 against the accGPT chatbot built upon GPT 3.5-Turbo and enhanced with knowledge of the ACR appropriateness criteria using a vectorized knowledge index.

## Methods

### Technical implementation of the chatbots

#### Data preparation and indexing

To develop and evaluate the proposed accGPT, we relied on the comprehensive collection of

209 topics with conditions potentially requiring diagnostic procedures from the ACR Appropriateness Criteria as foundational knowledge base (a schematic is presented in Figure 1).

**Figure 1.**
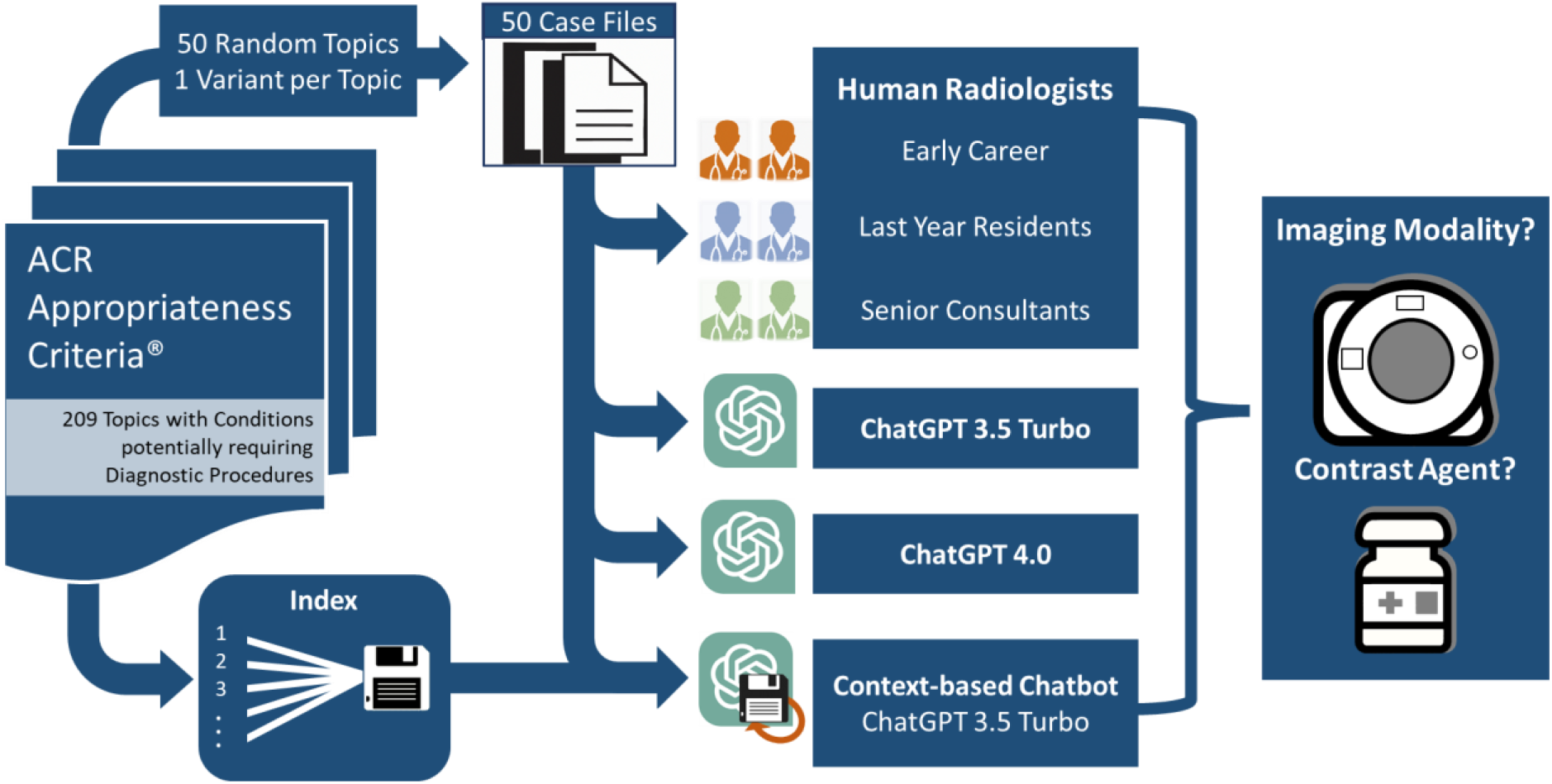
Schematic of the workflow of the case file creation, indexing of the context-based chatbot and performance analysis.

We employed LlamaIndex as an interface between external data and the GPT 3.5-Turbo (https://github.com/jerryjliu/llama_index; version 0.5.0). Text information was extracted from the ACR guidelines and indices were created using the GPTSimpleVectorIndex of LlamaIndex. For this, the document texts were divided into smaller chunks of up to 512 tokens (a measure of text content) and converted to data nodes. These nodes were then encoded using an embedding model (text-embedding-ada-002-v2 by OpenAI) and the result was stored in a dictionary-like structure.

#### Prompting strategy and answer synthesis

To customize the output of all chatbots for the case-based scenarios, the question posed to the system in each case was: “*Is imaging typically appropriate for this case? If so, please specify the most suitable imaging modality and whether a contrast agent is required. Exclude ‘May Be Appropriate’ and ‘Usually Not Appropriate’ as potential responses*.*”*

For GPT 3.5-Turbo and GPT 4, the direct output of was captured as response. For our context-based chatbot, the three best matching data nodes from the index were retrieved based on the embedding of the prompt. These nodes were used in a multistep answer creation and refinement method using GPT 3.5-Turbo and the final output was then captured.

### Preparation of case files

We sought to test the chatbot’s accuracy in comparison to human performance in a scenario resembling clinical routine. For this, 50 clinical case files were created based on the ACR appropriateness criteria. From the 209 topics with clinical conditions potentially requiring diagnostic procedures, 50 were randomly chosen. Subsequently, in each of those, one variant was randomly selected. Based on those 50 clinical conditions, case files were created resembling clinical routine referral notes by an experienced radiologist not involved in the reading. Clinical files included information on age, sex, chief complaint, past medical history and results from clinical testing while excluding the suspected pathology in most cases. The case files encompassed a wide range of topics and medical, some of which are rarely encounterd in a radiologst’s routineclinical practice (please see Supplementary Material for more details).

### Assessment of human and chatbot performance

The 50 case files were presented to six radiologists at different experience levels: two early career radiologists (first and second year of training), two advanced residents in their last year of training and two board-certified radiologists (with 11 and 12 years of experience). For each case file, appropriateness of imaging and if required, the most appropriate imaging modality and need for contrast agent administration was evaluated by all radiologists independently. During this assessment, no consultation of colleagues or guidelines was allowed.

We utilized a script-based approach on the 50 case files to perform a sixfold repetition testing for all three chatbots and assess their performance.

### Accuracy and agreement of radiologists and chatbots

The respective human- and chatbot-derived recommendations for imaging in the 50 case files were evaluated regarding their appropriateness according to the ACR guidelines, i.e. whether they met “usually appropriate” or “may be appropriate” criteria. Agreement of recommendations was evaluated for the six radiologists as well as the six outputs from the respective chatbots via Fleiss’ Kappa (R version 4.2.1, https://www.R-project.org/).

### Assessment of cost-effectiveness of radiologists and chatbots

Further, time to decision was assessed per reader to assess cost-effectiveness. Calculation of radiologists’s costs was based on the publicly available salary information for medical doctors working in university hospitals in Germany (https://oeffentlicher-dienst.info/aerzte/uniklinik/).

The costs using the GPT models were monitored on the billing output of the OpenAI webpage (https://platform.openai.com/account/usage) and validated with the token usage during the first run and price list.

## Code availability

The code for recreating the presented accGPT will be made available as an interactive interface using Gradio (https://gradio.app/) on an open Git repository as a jupyter notebook. Further code used in our analysis is available from the corresponding author upon reasonable request.

## Results

### Human and Chatbot Performance

In summary, chatbots performed at least at human performance level as given in Table 1 and Figure 2.

**Table 1.**
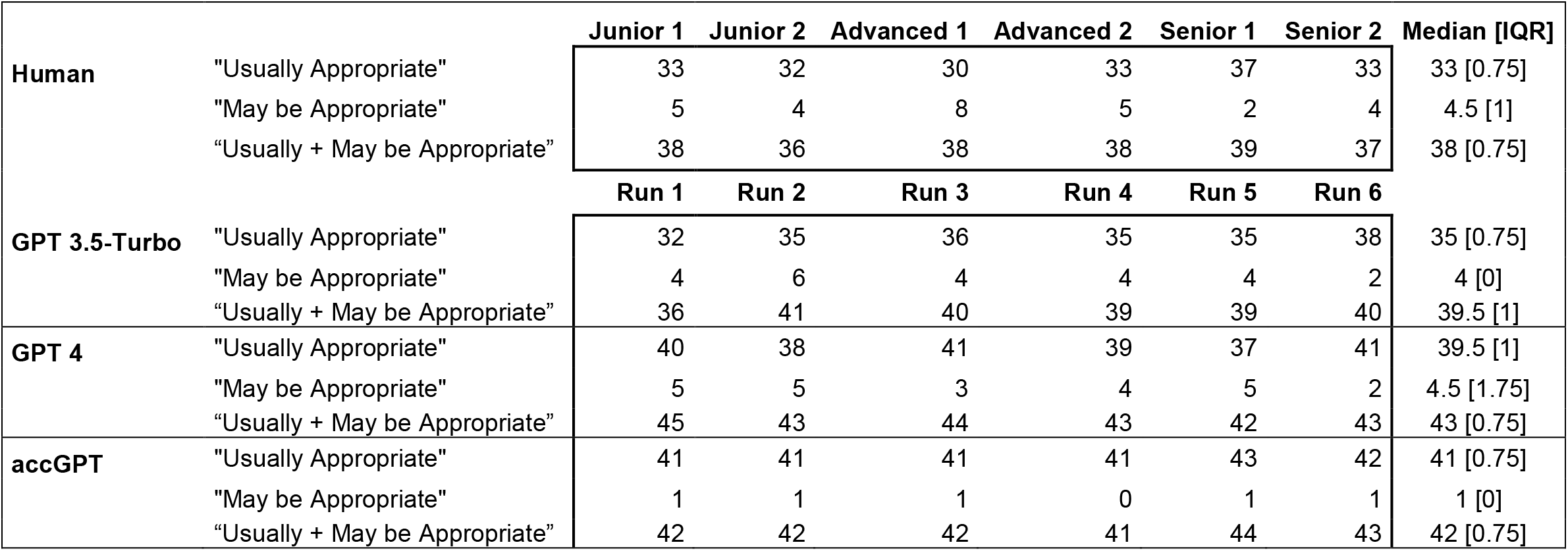
Correct answers on the case files according to the ACR appropriateness criteria

|  |  | Junior 1 | Junior 2 | Advanced 1 | Advanced 2 | Senior 1 | Senior 2 | Median [IQR] |
| --- | --- | --- | --- | --- | --- | --- | --- | --- |
| Human | "Usually Appropriate" | 33 | 32 | 30 | 33 | 37 | 33 | 33 [0.75] |
|  | "May be Appropriate" | 5 | 4 | 8 | 5 | 2 | 4 | 4.5 [1] |
|  | "Usually + May be Appropriate" | 38 | 36 | 38 | 38 | 39 | 37 | 38 [0.75] |
|  |  | Run 1 | Run 2 | Run 3 | Run 4 | Run 5 | Run 6 |  |
| GPT 3.5-Turbo | "Usually Appropriate" | 32 | 35 | 36 | 35 | 35 | 38 | 35 [0.75] |
|  | "May be Appropriate" | 4 | 6 | 4 | 4 | 4 | 2 | 4 [0] |
|  | "Usually + May be Appropriate" | 36 | 41 | 40 | 39 | 39 | 40 | 39.5 [1] |
| GPT 4 | "Usually Appropriate" | 40 | 38 | 41 | 39 | 37 | 41 | 39.5 [1] |
|  | "May be Appropriate" | 5 | 5 | 3 | 4 | 5 | 2 | 4.5 [1.75] |
|  | "Usually + May be Appropriate" | 45 | 43 | 44 | 43 | 42 | 43 | 43 [0.75] |
| accGPT | "Usually Appropriate" | 41 | 41 | 41 | 41 | 43 | 42 | 41 [0.75] |
|  | "May be Appropriate" | 1 | 1 | 1 | 0 | 1 | 1 | 1 [0] |
|  | "Usually + May be Appropriate" | 42 | 42 | 42 | 41 | 44 | 43 | 42 [0.75] |

**Figure 2.**
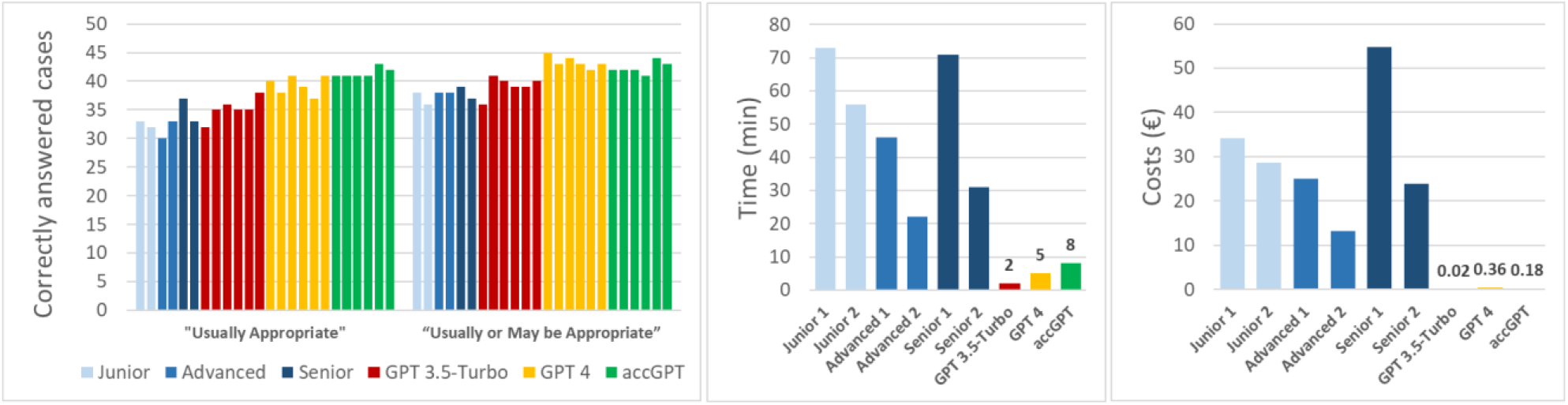
Comparison of the radiologists’ and chatbots’ performance regarding A) correctly provided recommendations in the 50 presented case files; B) time required to process the 50 cases; C) costs (€) incurred during the processing of the cases.

Radiologist’s recommendations for imaging in the 50 case files showed a median of 66% (95% CI: 62-70) of the answers meeting the “usually appropriate” criteria and a median of 9% (95% CI: 6-13) fulfilling the “may be appropriate” criteria. This results in a median of 76% (95% CI: 74-77) of the answers recommending either “usually appropriate” or “may be appropriate” imaging. Split into the different experience levels, we did not observe a robust difference in correct answers.

The six repetitive runs of the respective chatbot revealed higher proportions of “usually appropriate” answers for accGPT (median 83% (95% CI: 82-84)) compared to both the GPT 3.5-Turbo (median 70% (95% CI: 67-73)) and GPT 4 (median of 79% (95% CI: 76-81)). Furthermore, generic GPT 3.5-Turbo and GPT 4 provided a higher count of “may be appropriate” recommendations (median of 8% (95% CI: 6-10) and 9% (95% CI: 6-10), respectively) compared to accGPT (median 2% (95% CI: 1-2)). Case-based answer details are provided in Table 1.

### Consistency of radiologists and chatbots

The agreement for the “usually appropriate” answers among radiologists was moderate (0.44, 95% CI 0.36-0.51), while GPT 3.5-Turbo (0.62, 95% CI 0.55-0.69) and GPT4 (0.76, 95% CI

0.69-0.83) reached substantial agreement and accGPT reached almost perfect agreement (0.82, 95% CI 0.74-0.89). Agreement on the combined “usually + may be appropriate” responses followed a similar pattern with moderate agreement for radiologists (0.43, 95% CI 0.35 - 0.50), substantial agreement for GPT 3.5-Turbo (0.63; 95% CI 0.56 - 0.71) and almost perfect agreement for GPT 4 (0.84; 95% CI 0.77-0.91) and accGPT (0.83; 95% CI 0.75-0.87).

### Analysis on cost-effectiveness of radiologists and chatbots

The radiologists spend varying amounts of time evaluating the case files, with a mean duration of 49:48 min (SD 19 min; range 22-73 min). This resulted in a mean cost of 29.99€ (SD 12.78€, range 13.2-54.86€). GPT 3.5-Turbo required 2 min with 0.02€ costs, GPT 4 took 5 min and cost 0.36€, while accGPT needed 8 min and the token costs amounted to 0.18€. Overall, time and costs were significantly lower for the chatbots compared to the radiologists (both p=0.003).

## Discussion

Our results demonstrate the potential of the context-based accGPT chatbot in making imaging recommendations based on the ACR guidelines as it accepts standard clinical referral notes and provides concise recommendations on imaging in an end-to-end solution. Upon comparison against radiologists with varying levels of experience and two publicly available generic chatbots (GPT 3.5-Turbo and 4.0), we noted a superior accuracy and higher consistency in meeting the ACR recommendations. In addition, all chatbots were substantially more time efficient and less expensive than human radiologists in the evaluation of the case files.

The topic of our study is of great relevance as the insufficient usage of guidelines results in a large amount of inappropriate imaging ^3,6,7^. Inappropriate imaging is associated with increased costs for the healthcare systems, prolonged waiting lists, incorrect or delayed diagnoses, and potentially unnecessary exposure to ionizing radiation ^9^.

Notably, all evaluated chatbots achieved at least a human-level of performance, with accGPT providing consistently the highest proportion of “usually appropriate” recommendations. It is worth emphasizing that accGPT provided “may be appropriate” recommendations less frequently than human raters and the other chatbots, which reflects the quality of the recommendations. The significant improvement in performance from GPT version 3.5 to 4.0 is corroborated by other studies focusing on the simplification of radiological reports ^20^ and passing USMLE questions ^17^. Thus, it is reasonable to anticipate a substantial performance increase in a context-based chatbot utilizing GPT 4. In addition, the early stages of development for a dedicated combination of commercially available vector stores and up-to-date OpenAI models are underway in a limited alpha phase (https://openai.com/blog/chatgpt-plugins).

Nevertheless, ChatGPT itself was not explicitly designed for clinical decision making. This might result in incorrect recommendations and induce the “out-of-vocabulary” problem, potentially yielding outputs that are not entirely accurate or consistent with reality. However, incorporating specialized context-based knowledge, such as the ACR guidelines, addresses this constraint. This adaptability also enables the chatbot’s knowledge base to be updated or supplemented with additional guidelines or recommendations from other reputable sources. This also addresses concerns related to fake news and false data to ensure the reliability of the chatbot’s output ^22,24^.

For both the radiologists and the chatbots, we observed challenges particularly in the cases in which no imaging was appropriate (for example, case files 7, 27, 41). Rao et al. noted this limitation, too, as in their study ChatGTP recommended imaging in the only case where it was not necessary ^23^.

Since the ACR appropriateness criteria mainly comprise clinical conditions that require some kind of imaging, further investigation including more case vignettes without the need for imaging (e.g. dermatological pathologies) is of interest.

Existing tools that are based solely on trusted sources, such as the ESRiGuide have the drawback that clinical information from case vignettes need to be simplified for the input. This precludes assessing information from previous examinations and diagnostic workups ^13^. In contrast, the investigated chatbots allow for the direct input of the clinicians referral text. Furthermore, ChatGPT-based approaches allow for a deeper insight into the decision-finding process, as recently shown ^17^. This is also an advantage over less advanced natural language processing approaches that have already been developed as clinical decision support systems25.

Another benefit of the proposed approach is the observed reduction in evaluation duration as well as the marked cost efficiency of the algorithms compared to radiologists. From a technical standpoint, the integration into clinical information systems or radiological appointment scheduling systems is feasible. Considering the ever-increasing workload of radiologists ^26,27^, integrated tools like accGPT might constitute a relief in clinical routine.

When introducing AI-based solutions like accGPT into healthcare, addressing ethical, legal, and data security considerations is crucial. Additionally, the use of AI in clinical decision-making raises concerns about transparency, accountability, and potential biases ^22,28^. In the case of content-based chatbots, the usage as decision support tool, with an additional step of displaying the references can enhance transparency and trust.

Further optimization of the proposed accGPT should aim to include detailed assessment of the costs, availability, and potential radiation dose.

In summary, ChatGPT-based algorithms have the potential to substantially improve the decision-making for clinical imaging in accordance with ACR guidelines as decision-support tools. Specifically, a context-based algorithm was superior to the genetic chatbots, demonstrating the value of tailoring AI solutions to specific healthcare applications.

## Notes

### Competing Interest Statement

The authors have declared no competing interest.

### Funding Statement

This study did not receive any funding

